# Factors associated with secondhand smoke exposure among non-smoking employees in the workplace: A cross-sectional study in Qingdao, China

**DOI:** 10.1101/2022.01.28.22269994

**Authors:** Xiaocen Jia, Rui Wang, Xiaofei Qiu, Yiqing Huang, Yani Wang, Xiaorong Jia, Shanpeng Li, Yibo Wu, Fei Qi

## Abstract

**Objective:** The study was designed to describe secondhand smoke (SHS) exposure among non-smoking employees in the workplace and identify factors that related exposure in Qingdao.

**Methods:** The study subjects covered the key non-smoking places stipulated in the “Qingdao City Smoking Control Regulations”. Airborne nicotine concentration in the workplace and saliva cotinine concentration of employees were measured. Questionnaire included employees’ demographics factors, smoke-free measures in the workplace, the employer’s tobacco hazard knowledge and attitudes towards smoke-free policy.

**Results:** A total of 222 non-smoking employees and 46 employees were included in the study. The median concentrations of airborne nicotine and salivary cotinine were 0.389 μg/m^3^ and 0.575 ng/ml. Educational status, average number of smokers per day and exposure time of SHS in the workplace, whether to divide smoking and non-smoking areas were related to the airborne nicotine concentration significantly. Age, educational status, exposure time of SHS in the workplace, tobacco control training and publicity and whether the employers support the “Qingdao Tobacco Control Regulation” were related to the salivary cotinine concentration significantly.

**Conclusions:** Exposure to SHS is highly prevalent among non-smoking employees in the workplace. Interventions to reduce SHS exposure in the workplace are urgently needed.

## Introduction

Secondhand smoke (SHS), also referred to as environmental tobacco smoke, is the combination of smoke exhaled by smokers and the smoke from the burning tip of the cigarette[1]. SHS is a serious health hazard that can cause or worsen a wide range of health effects, including respiratory and cardiovascular diseases[2]. There is no risk-free level of secondhand smoke exposure and even brief exposure can be harmful to health[3]. Comprehensive smoke-free policies have been successful in protecting those who do not smoke, and are the only way to fully protect their health[3]. Therefore, the World Health Organization (WHO) recommends the adoption and implementation of comprehensive national smoke-free legislation to fully protect people from SHS.

An increasing number of countries have enacted partial or comprehensive national smoke-free laws, which generally prohibit smoking in indoor workplaces, indoor public places, public transportation and, as appropriate, other public places as required by the World Health Organization Framework Convention on Tobacco Control[4]. However, 50.9% of adults working indoors (216.9 million adults) were exposed to SHS in the workplace according to the 2018 Global Adult Tobacco Survey. A study from the United States showed that 8.6% of nonsmoking workers reported exposure to SHS frequently in the workplace even in states with smoke-free laws in worksites[5]. The workplace is still the source of most SHS exposure for non-smoking adults[6]. Exposure to SHS in the workplace has been recognized as one of the major occupational hazards leading to the prevalence of occupational cancer among non-smokers[7]. In addition, the workplace is one of the settings where a large number of deaths related to exposure to SHS have been reported. The International Labor Organization estimates that approximately 14%, about 200,000, of all work-related deaths due to diseases are related to exposure to SHS in the workplace around the world[8].

China is home to the largest number of smokers in the world[2]. It is also estimated that 70% of Chinese adults are frequently exposed to SHS[9]. In recent years, local-level tobacco control policy initiatives have emerged in China. All tier 1 cities, namely Beijing, Shanghai and Shenzhen have implemented local smoke-free policies. Qingdao, a new first-tier city (a tier immediately below tier 1) in eastern Shandong province, enacted the smoke-free law on 31 Aug 2013 that prohibits smoking in indoor workplaces. The results of the 2014 Qingdao Adult Tobacco Epidemic Survey showed that the exposure to second-hand smoke in Qingdao is relatively serious in the early stage of smoke-free law and the overall exposure rate of second-hand smoke in indoor workplaces is 32.7%. The workplace was observed to be a strategic place for SHS exposure as most adults spend more than half a day at their workplace[10]. However, self-reporting was usually used to assess the exposure of employers in the workplace in the past, but self-reporting may underestimate or overestimate the actual exposure. This may be due to a lack of knowledge about how SHS is distributed in the workplace or inaccurate employee reports, indicating that objective measures are more reliable[11]. The air nicotine concentration can be measured to assess SHS exposure in a specific environment[12]. Nicotine is specific to tobacco smoke, sensitive at low concentrations, and easy to collect[13; 14]. Nicotine is often used to evaluate SHS in different indoor settings such as homes or workplaces such as hospitality venues[13; 15; 16], and it is also considered a reliable environmental gold standard[17]. The biomarker cotinine, a metabolite of nicotine, can be used to determine individual SHS exposure, which reveals the nicotine intake of adults in the past 24 -48 hours[18]. In addition, saliva is a suitable surrogate diagnostic tool for other body fluids as saliva testing is cost-effective, simple and non-invasive[19]. In individuals, salivary cotinine concentrations have been shown to be approximately the same as plasma concentrations[20].

Until now, only few studies aimed to focus on the determinants of SHS exposure, although this information is needed for adequate public health policies to protect non-smokers. Studies on the determinants of SHS exposure in China are scarce. Therefore, the aim of this study is to describe the exposure to SHS of non-smoking employees in the workplace. SHS exposure of each non-smoking employee is described using a seven-day nicotine accumulation measurement and a measure of salivary cotinine. A secondary aim is to explore the relationship between airborne nicotine concentrations in the workplace and the employees’ salivary cotinine level with reported employees’ demographics factors, smoke-free measures in the workplace and employer’s tobacco hazard knowledge and attitudes towards smoke-free policy. The information provided in this study will allow for the development and implementation of targeted preventive measures for the reduction of SHS exposure.

## Methods

### Study design

The study subjects covered the key non-smoking places stipulated in the “Qingdao City Smoking Control Regulations”. Convenience sampling methods were used to select a total of 46 workplaces, including three categories: restaurants, bars, office buildings. The inclusion criteria of the subjects were informed consent, no smoking (never smoked or quit smoking for more than half a year), age over 18 years old, working time in the workplace for 3 hours or more on the test day, exposure time to SHS in non-workplaces (such as homes, other public places) is less than 1 hour on the test day. The exclusion criteria were those who were unwilling to participate in this research or unwilling to cooperate, and those who were unable to communicate normally, such as text dyslexia. The employers and employees completed a questionnaire respectively to determine the employees’ demographics factors, smoking ban measures in the workplace and the employer’s knowledge of tobacco hazards. Measurements included airborne nicotine concentration in the workplace and saliva cotinine concentration of employees. The survey instruments, protocols and the process for obtaining the informed consent for participants were carried out in accordance with relevant guidelines and regulations and were approved by the Institutional Review Board of Qingdao Municipal CDC.

### Airborne Nicotine Sampling

We collected airborne nicotine using a passive sampling device that contained a 37 mm diameter filter treated with sodium bisulfate. According to “Technical Specification for Sampling of Nicotine Passive Sampler in Ambient Air”, the filter membrane is processed and the sampler is installed. The sampler is hung at a distance of 1.5 to 2 meters from the ground and avoids places with no air circulation. After a seven-day sampling period, nicotine was enriched on the absorption membrane, and finally the absorption membrane samples were detected in the analytical laboratory. The laboratory is certified by the Johns Hopkins University Global Tobacco Control Institute for laboratory testing capabilities for nicotine content in air samples. The total amount of nicotine absorbed by each filter was quantified by gas chromatography combined with mass spectrometry. Nicotine concentration is calculated by dividing the total amount of nicotine by the rate of air flow and the length of time (in minutes) for which the device was installed[15]. The analysis procedure is certified by the ISO-17025, and has a nicotine limit of detection (LOD) of 0.02 μg/m^3^ for 1 week of exposure.

### Saliva Sample Collection

Before sample collection, the researchers wiped their hands thoroughly with baby wipes to minimize the chance of contamination. The saliva sample is collected with a professional saliva collection tube. The tested personnel shall chew the cotton swab for 45 seconds, then put the chewed cotton swab back into the saliva collection tube, cover the sealing cover and centrifuge to collect saliva. The collected saliva samples need to be transported to the CDC on the same day for cryopreservation at -20°C, and sent to the laboratory for cotinine concentration detection within 7 days after collection. The lower limit of saliva cotinine is 0.1 ng/mL. Cotinine concentrations below the limit of quantification were designated as half the level of quantification (0.05 ng/mL).

### Study variables

The study is exploratory. In the multiple linear regression model, the dependent variables are airborne nicotine concentration in the workplace and salivary cotinine concentration among employees. The information on the independent variable was assessed by the questionnaire survey. The first model aims to assess the demographics factors being related to SHS exposure. The independent variables included information on sex (“male” and “female”), age groups (“18-30 years”, “31-45 years”, “46-60 years”), educational status (“low”, “middle” and “high”), average number of smokers per day in the workplace (“<1”, “1∼10”, “>10”), exposure time of SHS in the workplace (“<1 h”, “1∼6h”, “>6 h”) and colleague smoking (“yes” and “no”). The second model aims to assess the smoke-free measures being related to SHS exposure in the workplace. The independent variables included information on indoors smoking bans (“yes” and “no”), whether to divide smoking and non-smoking areas (“yes” and “no”), regulations on tobacco control (“yes” and “no”), leaders in charge of tobacco control (“yes” and “no”), tobacco control supervisor (“yes” and “no”), tobacco control training and publicity (“yes” and “no”), employees discourage smoking actively (“yes” and “no”). The third model aims to assess the relationship between employer’s knowledge and attitudes towards smoking bans and SHS exposure in the workplace. The independent variables included information on awareness of whether to support the “Qingdao Tobacco Control Regulation”(“yes” and “no”), the effect of the “Qingdao Tobacco Control Regulation” on the workplace (“beneficial”, “unhelpful” and “no effect”), awareness of the “Qingdao Tobacco Control Regulation”(“yes” and “no”), awareness of the maximum fines of the “Qingdao Tobacco Control Regulations” (“yes” and “no”), and employers’ knowledge of tobacco hazard score.

### Data analysis

SPSS 26.0 was used for statistical analyses in this study. The significance level was set at 0.05. First, we checked the distribution of the airborne nicotine concentration and salivary cotinine concentration, and found they were in non-normal distribution. Second, logarithmic transformation of skewness data was performed to provide an approximate normal distribution for statistical analysis. Correlations between airborne nicotine concentration and salivary cotinine concentration were tested on log - transformed data using a Pearson correlation. A multiple linear regression analysis was used to determine whether any factors were significantly related to airborne nicotine and salivary cotinine concentration. Participants with missing data were excluded only from the factor analysis associated with the missing variable.

## Results

### Participant demographics, Airborne nicotine and Salivary Cotinine Levels

Table 1 shows the demographics of all participants and the airborne nicotine and salivary cotinine levels within each demographic group. 250 non-smoking employees and 46 employers from restaurants, bars and office buildings participated in the study, and data related to saliva cotinine concentration were obtained from 235 employees. In addition, to control for misreported smoking status, we excluded 13 subjects with saliva concentrations exceeding 15 ng/ml. A total of 222 employees were included in the study. 32 employees had values below the detection limit(<0.1ng/ml), giving a value of 0.05 ng/ml for analysis. The median concentrations of airborne nicotine and salivary cotinine were 0.389 μg/m^3^ and 0.575 ng/ml and there was a significant positive correlation of 0.382 (p<0.01) between them.

**Table 1.** Summary of the demographic variables.

| **Demographic** | **Employee**<br>**n(%)** | **Employer**<br>**n(%)** | **Airborne**<br>**nicotine**<br>**concentration**<br>**($\mu\text{g}/\text{m}^3$)** | **Salivary**<br>**cotinine**<br>**concentration**<br>**(ng/ml)** |
| --- | --- | --- | --- | --- |
| **Gender** |  |  |  |  |
| **male** | 78(35.1) | 33 (71.7) | 0.381 | 0.649 |
| **female** | 144(64.9) | 13 (28.3) | 0.389 | 0.498 |
| **Age** |  |  |  |  |
| **18-30** | 120(54.1) | 20 (43.5) | 0.389 | 0.641 |
| **31-45** | 92(41.4) | 13 (28.3) | 0.385 | 0.294 |
| **46-60** | 10(4.5) | 13 (28.3) | 0.415 | 1.135 |
| **Educational status** |  |  |  |  |
| **Low** | 27 (12.2) | 4 (8.7) | 0.623 | 1.048 |
| **Middle** | 75 (33.8) | 16(34.8) | 0.623 | 0.839 |
| **High** | 120 (54.1) | 26 (56.5) | 0.306 | 0.317 |
| **Type of workplace** |  |  |  |  |
| **Restaurants** | 88(39.6) | 21(45.7) | 0.914 | 1.165 |
| **Bars** | 44(19.8) | 10(32.6) | 0.166 | 0.395 |
| **Office buildings** | 90(40.5) | 15(21.7) | 0.328 | 0.210 |
| **Average number of smokers per day in the workplace** |  |  |  |  |
| **<1** | 131(59.0) |  | 0.284 | 0.412 |
| **1~10** | 71(32.0) |  | 0.604 | 0.858 |
| **>10** | 20(9.0) |  | 0.905 | 0.864 |
| **Exposure time of SHS in the workplace** |  |  |  |  |
| **<1 h** | 155(65.8) |  | 0.306 | 0.355 |
| **1~6h** | 65(27.9) |  | 0.666 | 0.930 |
| **>6h** | 15(6.3) |  | 1.055 | 1.311 |
| **Colleague smoking** |  |  |  |  |
| **Yes** | 28(12.6) |  | 0.385 | 0.504 |
| **No** | 194(87.4) |  | 0.389 | 0.575 |
| **Total** | 222 (100) | 46 (100) | 0.389 | 0.575 |

### Relationship between employees’ demographics factors and SHS exposure in the workplace

In order to explore the relationship between employees’ demographics factors and airborne nicotine, saliva cotinine, a multiple linear regression analysis was performed (see Table 2). The results show that educational status of employees, average number of smokers per day in the workplace, exposure time of SHS in the workplace were related to the airborne nicotine concentration significantly. According to these results, higher educational status of employees is associated with lower air nicotine concentration in the workplace (β=-0.286; 95%CI: -0.663, -0.105). The average number of smokers per day in the workplace is related to the air nicotine concentration significantly (p=0.041). The more the number of smokers per day in the workplace, the higher the air nicotine concentration (β=0.141; 95%CI:0.003, 0.401). The longer exposure time of SHS was associated with higher airborne nicotine concentration (p=0.008). Compared with the SHS exposure time of employees in the workplace<1h, the SHS exposure time of employees>6h showed an β of 0.193 (95 % CI: 0.143, 0.918). In terms of salivary cotinine, age and educational status of employees, exposure time of SHS in the workplace were significant predictors of salivary cotinine concentration. Employees aged 36-45 years showed an β of -0.153 (95 % CI: -0.408, -0.034) compared to employees aged 18-35 years. With increasing age the likelihood of SHS exposure decreased. Consistent with the result of nicotine concentrations in the workplace, higher educational status is associated with lower salivary cotinine concentration among employees (β=-0.241; 95%CI: -0.637, -0.052). The exposure time of SHS in the workplace is related to the saliva cotinine concentration among employees. Compared with the SHS exposure time of employees in the workplace <1 h, the SHS exposure time of employees1-6 h and > 6h showed an β of 0.257 (95 % CI:0.184, 0.631) and 0.217 (95 % CI:0.229, 1.042) respectively.

**Table 2.**
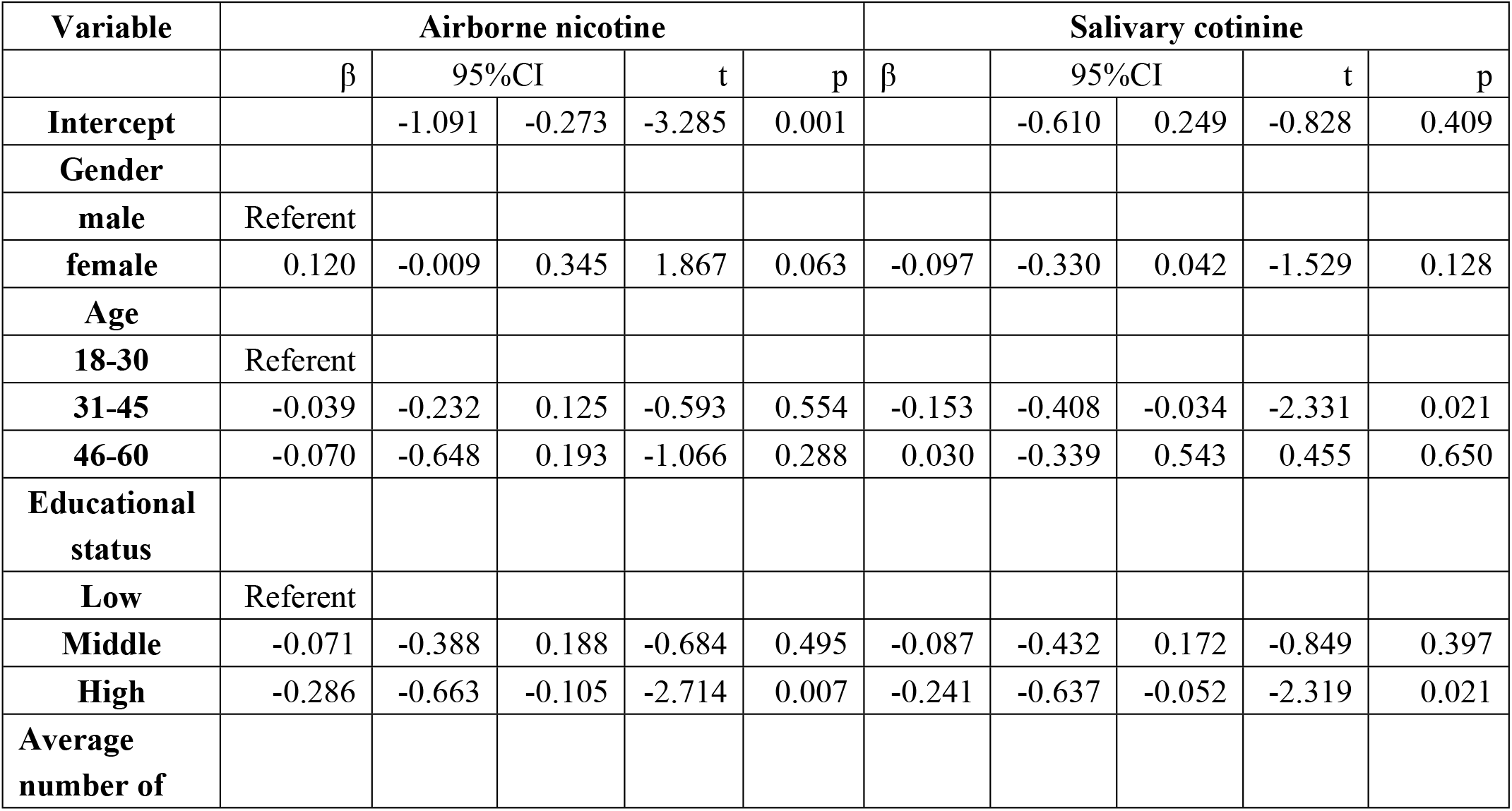
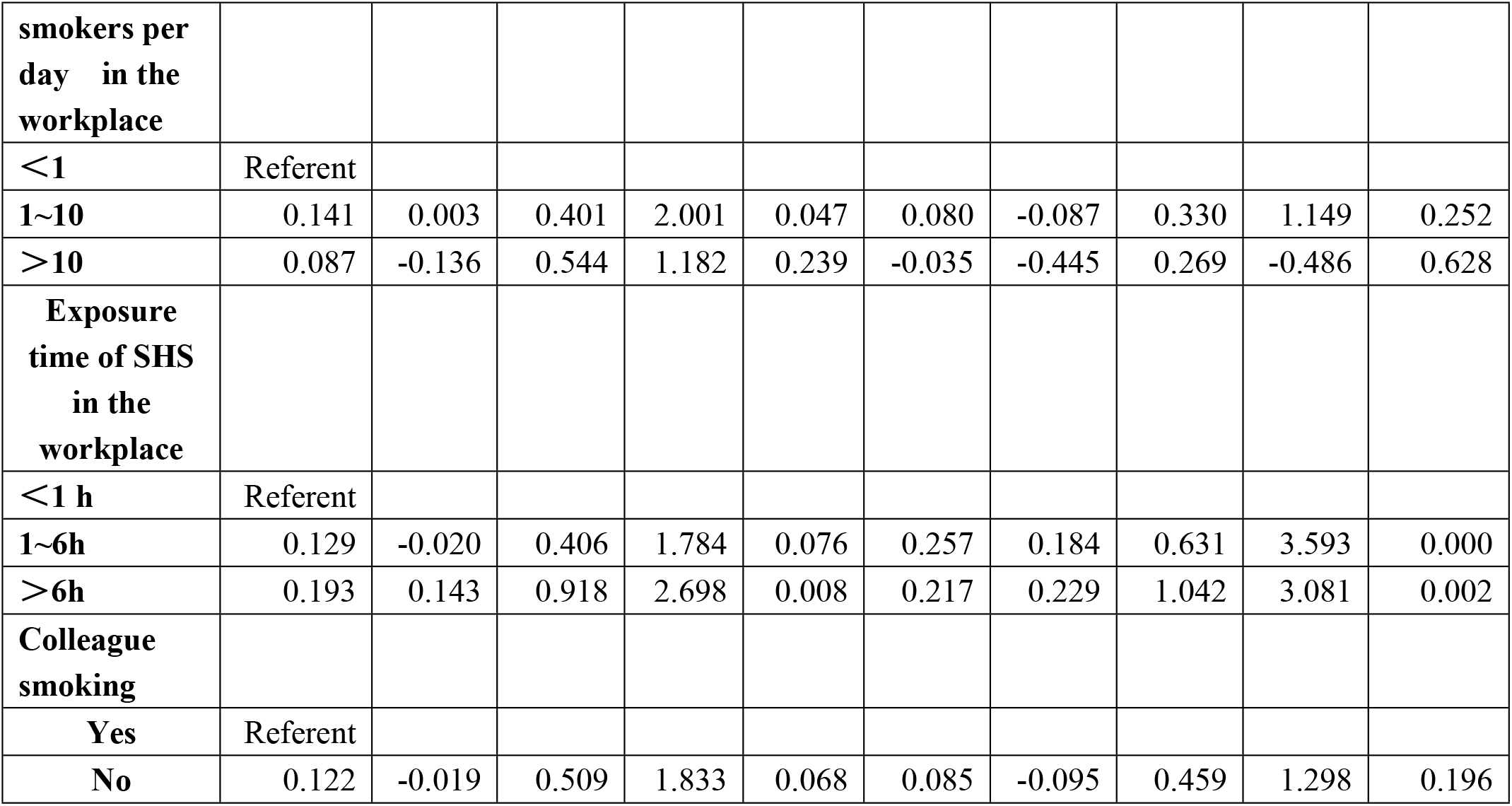
Multivariate linear regression analysis of employees’ demographics factors associated with airborne nicotine concentration and salivary cotinine concentration (n = 222).

### Relationship between smoke-free measures and SHS exposure in the workplace

After controlling for employees’ demographic factors such as gender and age, relationship between smoke-free measures in workplaces and airborne nicotine, saliva cotinine is shown in Table 3. Whether to divide smoking and non-smoking areas in the workplace was significant predictors of airborne nicotine. Compared with the division of smoking and non-smoking area in the workplace, the airborne nicotine concentration is higher without division (β=0.193; 95%CI:0.079, 0.482). Tobacco control training and publicity in the workplace was a significant predictor of employees’ salivary cotinine concentration (p=0.045). Compared with employees who received tobacco control training and publicity in the workplace, employees who did not receive had higher airborne nicotine concentrations in the workplace. (β=0.179; 95%CI:0.006, 0.566).

**Table 3.** Multivariate linear regression analysis of smoke-free measures in workplaces associated with airborne nicotine concentration and salivary cotinine Concentration (N = 222).

| Variable | Airborne nicotine |  |  |  |  | Salivary cotinine |  |  |  |  |
| --- | --- | --- | --- | --- | --- | --- | --- | --- | --- | --- |
| | $\beta$ | 95%CI | | t | p | $\beta$ | 95%CI | | t | p |
| **Intercept** |  | -1.091 | -0.273 | -3.285 | 0.001 |  |  | -0.610 | 0.249 | -0.828 |
| **Indoor smoking bans** |  |  |  |  |  |  |  |  |  |  |
| **Yes** | Referent |  |  |  |  |  |  |  |  |  |
| **No** | 0.117 | -0.048 | 0.457 | 1.596 | 0.112 | 0.084 | -0.113 | 0.425 | 1.146 | 0.253 |
| **Whether to divide smoking and non-smoking areas** |  |  |  |  |  |  |  |  |  |  |
| **Yes** | Referent |  |  |  |  |  |  |  |  |  |
| **No** | 0.193 | 0.079 | 0.482 | 2.746 | 0.007 | 0.125 | -0.020 | 0.408 | 1.787 | 0.075 |
| **Regulations on tobacco control** |  |  |  |  |  |  |  |  |  |  |
| **Yes** | Referent |  |  |  |  |  |  |  |  |  |
| **No** | 0.027 | -0.235 | 0.33 | 0.333 | 0.739 | 0.071 | -0.169 | 0.432 | 0.865 | 0.388 |
| **Leaders in charge of tobacco control** |  |  |  |  |  |  |  |  |  |  |
| **Yes** | Referent |  |  |  |  |  |  |  |  |  |
| **No** | 0.103 | -0.218 | 0.518 | 0.804 | 0.423 | -0.101 | -0.547 | 0.236 | -0.782 | 0.435 |
| **Tobacco control supervisor** |  |  |  |  |  |  |  |  |  |  |
| **Yes** | Referent |  |  |  |  |  |  |  |  |  |
| **No** | -0.150 | -0.587 | 0.155 | -1.147 | 0.253 | -0.087 | -0.527 | 0.261 | -0.665 | 0.507 |
| **Tobacco control training and publicity** |  |  |  |  |  |  |  |  |  |  |
| **Yes** | Referent |  |  |  |  |  |  |  |  |  |
| **No** | 0.088 | -0.131 | 0.396 | 0.994 | 0.321 | 0.179 | 0.006 | 0.566 | 2.016 | 0.045 |
| **Employees discourage smoking actively** |  |  |  |  |  |  |  |  |  |  |
| **Yes** | Referent |  |  |  |  |  |  |  |  |  |
| **No** | -0.107 | -0.731 | 0.095 | -1.518 | 0.131 | -0.112 | -0.793 | 0.086 | -1.587 | 0.114 |

### Relationship between employer’s tobacco hazard knowledge and attitudes towards smoke-free policy and second-hand smoke (SHS) exposure of employees in the workplace

After controlling for employers’ demographic factors such as gender and age, relationship between employer’s tobacco hazard knowledge and attitudes towards smoke-free policy and airborne nicotine, saliva cotinine is shown in Table 4. Whether employers support the “Qingdao Tobacco Control Regulation” in the workplace is related to the saliva cotinine concentration of employees significantly. Compared with supporting the “Qingdao Tobacco Control Regulation”, employees in workplaces where employers do not support the “Qingdao Tobacco Control Regulation” have higher saliva cotinine concentration (β=0.359; 95%CI: 0.050, 1.843).

**Table 4.**
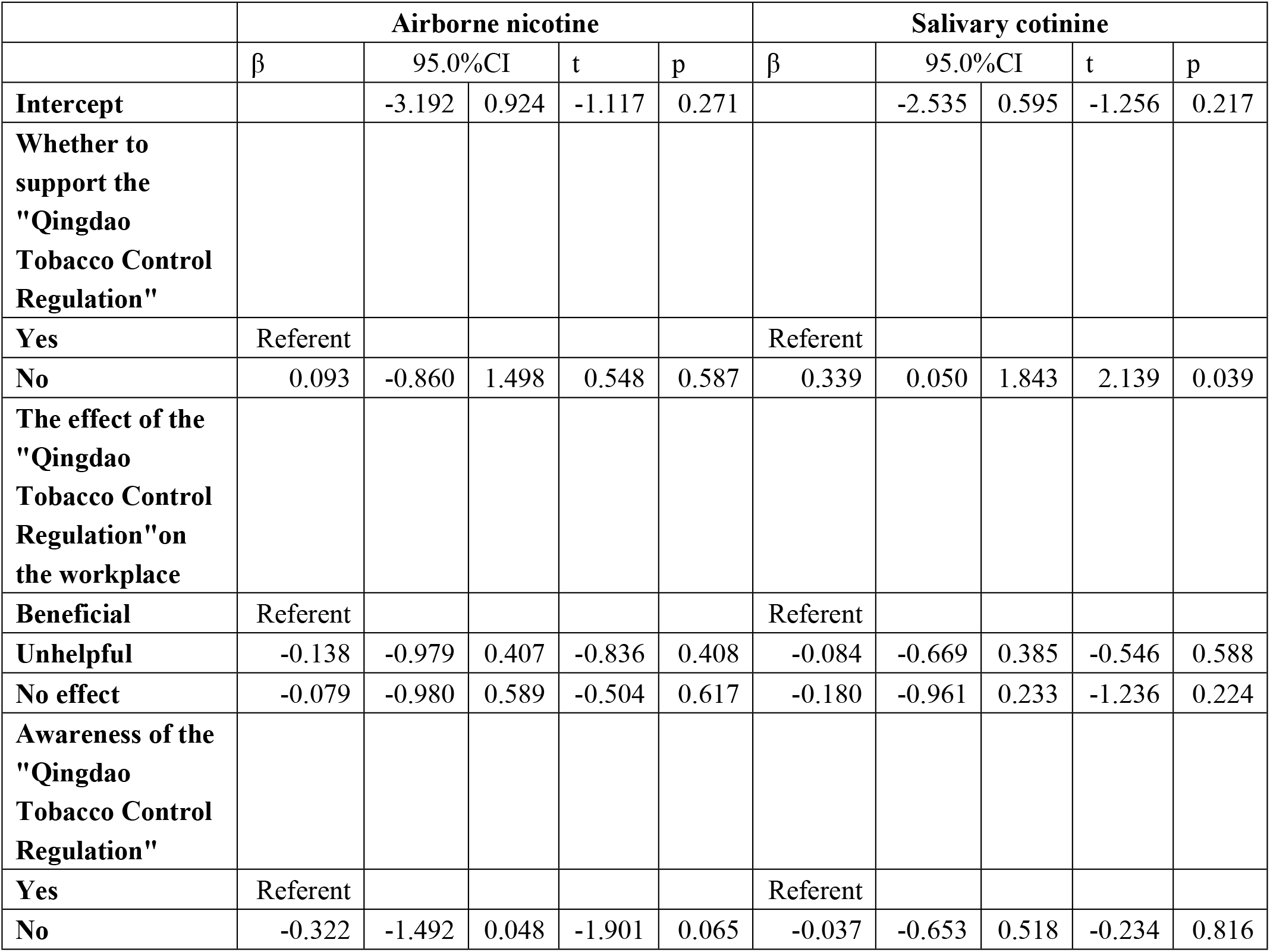
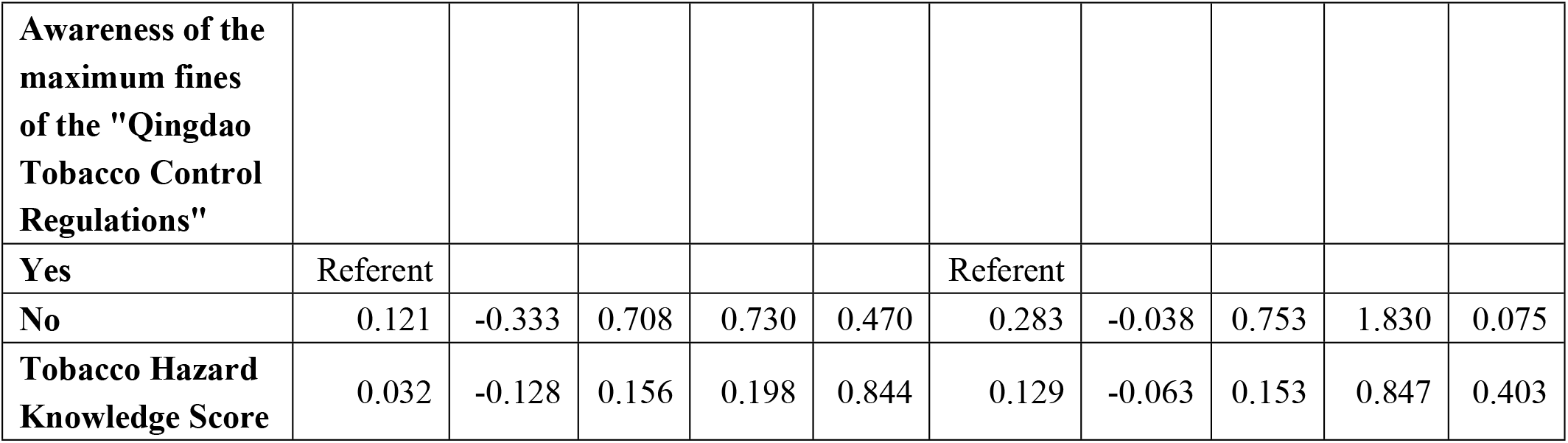
Multivariate Linear Regression Analysis of Employers’ Knowledge and Attitudes towards Smoking Bans Associated with Airborne Nicotine Concentration and Salivary Cotinine Concentration (N = 46).

## Discussion

Airborne nicotine has been widely used as a SHS indicator in occupational and non-occupational settings[13]. Measurements of airborne nicotine, a specific component of tobacco, reflect tobacco smoke exposure. The passive nicotine sampler is used to measure nicotine in the air with high sensitivity and specificity, and has been gradually applied in the evaluation of environmental tobacco smoke pollution recent years[21]. In this study, we found that the median concentrations of airborne nicotine was 0.389 μg/m^3^, lower than the partial monitoring results of indoor airborne nicotine in some workplaces in Qingdao in 2016[21], but the workplace remains an environment where further improvements can be made to reduce SHS exposure.

In terms of employees’ demographics, factors related to airborne nicotine concentrations include educational status, average number of smokers per day in the workplace and exposure time of SHS of employees in the workplace. The higher the airborne nicotine concentration in the workplace with lower level of education of people revealed in this study are comparable with previous literatures[22; 23]. Those of lower educational attainment and socioeconomic status are still less likely to be covered by smoke-free laws in office buildings, restaurants, and bars and more likely to be exposed to SHS[6]. In addition, less educated people have lower awareness of the health effects of smoking[24]. Thus evidence-based interventions and tailored strategies are warranted to reduce the exposure rate of SHS among employees in the workplace, with priority to low educational status groups. The more the number of smokers per day in the workplace, the higher the airborne nicotine concentration. Other studies also showed that nicotine concentrations increased with the number of cigarettes lit[25; 26], which is consistent with this study. Analysis of PM2.5 levels in smoking locations showed an increase of 129 ug/m^3^ in PM2.5 levels per smoker per 100 m3 room volume[27]. The PM2.5 concentration is positively correlated with the airborne nicotine concentration[28], so the airborne nicotine concentration will increase accordingly. The longer exposure time of SHS was associated with higher airborne nicotine concentration. SHS increases exposure to airborne nicotine directly, and when exposed to SHS, nonsmokers inhale 60% to 80% of airborne nicotine, absorbing concentrations similar to those absorbed by smokers, and showing high levels of nicotine biomarkers[29].

Regarding to smoke-free measures in the workplace, whether to divide smoking and non-smoking areas are important factors influencing airborne nicotine concentration. The results of this study show that workplaces with smoking and non-smoking areas tend to have lower airborne nicotine concentrations. However, a previous study showed that the geometric mean PM2.5 levels in non-smoking rooms is much higher than in completely smoke-free reception venues even if non-smoking and smoking areas were spatially separated into two rooms[27]. As early as 2006, a US Surgeon General’ report concluded that the scientific evidence consistently showed that mechanical systems and separated areas could not protect the population from SHS exposure[30]. This study only investigated whether the workplace is divided into smoking and non-smoking areas. The workplaces that are not divided may include completely smoke-free places and places that are not smoke-free, resulting in a certain deviation in the results.

Cotinine is the main metabolite of nicotine, and its concentration in body fluids is determined by nicotine metabolism rate and cotinine clearance. Although there may be individual differences in salivary cotinine concentration due to these parameters, it remains an important indicator of nicotine dependence[31]. The results of this study showed that the median saliva cotinine concentrations of employees in restaurants, bars and office buildings were respectively 1.165, 0.395, 0.210 ng/ml. Restaurants and bars are the most important source of SHS exposure for more than half of the non-smoking adults. For non-smoking servers living in smoke-free homes, time spent in restaurants and bars still dominates the total SHS exposure time[32]. Exposure to SHS in restaurants and bars alone has a much higher than acceptable health risk of developing asthma among customers and servers, as well as death from cancer and heart disease, a study has shown[33]. The results of this study suggest that age was a significant predictor of employee’s salivary cotinine, with younger employees having higher salivary cotinine levels. A study from Germany[23] has already highlighted higher SHS exposure among young people, which is also consistent with the results of this study. Educational status, exposure time of SHS are also important factors affecting the saliva cotinine among employees, which is consistent with the result of airborne nicotine concentration in the workplace.

Tobacco control training and publicity for employees and employer support for the “Qingdao Tobacco Control Regulation” in the workplace were also significant predictors of employee’s salivary cotinine. More training for employees may enhance their knowledge towards smoking hazards and improve employees’ support for tobacco control in the workplace[34], thereby promoting the implementation of smoke-free policy in the workplace. The most immediate authority at the workplace is the employers[35]. Employers support the “Qingdao Tobacco Control Regulation” in the workplace and are more inclined to devise plans to increase knowledge and attitude towards tobacco control of the employees through training program or an awareness campaign[10], reducing their exposure to second-hand smoke in the workplace, thereby reducing employees’ cotinine levels. On the contrary, employers’ poor attitudes toward the smoke-free policy resulted in poor action towards preventing SHS exposure.

The strong positive association between airborne nicotine and salivary cotinine validates the use of either measure as an index of employees’ SHS exposure in the workplace. Our findings suggest that smoke-free measures and employers’ attitudes toward tobacco control are very important in reducing SHS exposure in the workplace. Reducing SHS exposure in the workplace requires a joint effort by employers and employees.

### Limitations and Strengths

The study has some limitations which have to be acknowledged. Our analysis was limited to non-smoking employees, although smoking employees experienced negative health effects from SHS in addition to the effects of active smoking. Second, we included only exposure that occurred in the workplace, although people may also be exposed to SHS in other settings, such as home, parks, public buildings, and other venues. Moreover, the small sample size is also one of the limitations of this study, which limits further analysis of these data.

This study also had a number of strengths. Our measures of SHS exposure have advantages over some measures used in other studies. An important strength of this study is the assessment of SHS exposure measuring airborne nicotine concentrations, a specific tracer often used as a surrogate for other toxic and carcinogenic components in tobacco[36]. Measuring airborne nicotine concentrations allows us to precisely quantify secondhand smoke exposure levels and compare them to previous measurements in other countries. Measuring airborne nicotine concentrations allows us to precisely quantify SHS exposure levels and compare them to previous measures in other countries[37; 38]. The use of salivary cotinine in this study as a specific biomarker of SHS exposure in the past 2-5 days is another strength of this study. In addition, the analytical method for evaluating salivary cotinine is highly sensitive.

## Conclusion

Despite the implementation of Qingdao Smoking Control Regulations in 2013, the workplace remains an important place for second-hand smoke exposure. The ‘SHS issue’ has not yet been ‘solved’ and the public health community needs to continue their efforts and consider taking further measures to protect non-smokers from SHS. Therefore, not only must the legislation be implemented, but also further public health strategies must be considered to promote a total smoking ban in the workplace.

## Data Availability

All relevant data are within the manuscript and its Supporting Information files.

## Acknowledgments

We are grateful to thank Medical and Health Discipline Construction Project of Qingdao and iPhase Pharmaceutical Services for helping with this paper.

## Author Contributions

Conceptualization: Fei Qi.

Data curation: Xiaocen Jia, Rui Wang, Xiaofei Qiu, Yiqing Huang, Yani Wang, Xiaorong Jia, Shanpeng Li.

Formal analysis:Xiaocen Jia, Rui Wang, Xiaofei Qiu, Yiqing Huang.

Methodology: Xiaocen Jia, Rui Wang, Xiaofei Qiu, Yiqing Huang.

Software: Xiaocen Jia, Rui Wang, Xiaofei Qiu, Yiqing Huang.

Supervision: Fei Qi.

Writing – original draft: Xiaocen Jia.

Writing – review & editing: Xiaocen Jia, Yibo Wu.

